# Age Specific Months of Mortality (MOMa) from Endemic and Pandemic (Covid 19) diseases

**DOI:** 10.1101/2020.11.17.20233197

**Authors:** Christopher M Rembold

**Affiliations:** Cardiovascular Division, Department of Internal Medicine, University of Virginia Health System Charlottesville, Virginia 22908-0146 USA

**Keywords:** Risk, mortality, covid 19, weight, impact

## Abstract

People do not naturally understand risk. We fear things that happen rarely like kidnapping while ignoring common risks like motor vehicle crashes. We also do not fully comprehend the large effect that age has on risk. In this paper, I introduce a concept that I call age specific months of mortality, abbreviated MOMa, a statistic that will allow people to understand their risk of death within their age group. In a year without excess mortality, i.e. no pandemic, individual causes of death will add up to a total of 12 MOMa. Excess mortality, e.g. a pandemic, adds MOMa beyond 12. For people in their 20s, the MOMa is 5 for accidents, 1.9 for suicide, 1.6 for homicide, and 1.2 for Covid-19. For people in their 60s, the MOMa is 12 for Covid-19, 4 for cancer, 2.6 for coronary heart, and treatment of Covid-19 with dexamethasone reduces MOMa from 12 to 7 months.

## INTRODUCTION

People do not naturally understand risk. We fear things that happen rarely like kidnapping while ignoring common risks like motor vehicle crashes. We also do not fully comprehend the huge effect that age has on risk. In this paper, I introduce a concept that I call age specific months of mortality which I abbreviate MOMa. This idea was sparked by an interview of Professor David Spiegelhalter on the BBC radio/podcast program “More or Less” on 28 March 2020 (available on iTunes). First, I will describe the MOMa for various endemic diseases, then discuss the MOMa for excess mortality caused by the pandemic disease Covid-19, and finally describe calculation methods.

## RESULTS

For endemic diseases such as accidents and stroke, there is a predictable number of deaths yearly. If there is no excess mortality, i.e. no pandemic or war, the yearly number of total deaths is also predictable. If we normalize the total endemic deaths to the value 12, then there would be 12 months of total endemic deaths yearly. Specific causes of endemic death would be a fraction of total deaths. For example, if stroke were responsible for one out of six endemic deaths (18%), then stroke would cause 12 x (1/6) or 2 months of mortality.

Since deaths and causes of death are strongly dependent on age, it is appropriate to have the months of mortality statistic stratified by age, so I propose a statistic called age-specific months of mortality, abberviated MOMa. The left hand panel of Fig. 1 shows the MOMa for the leading causes of death by decile of age based on 2015 United States data from the Centers for Disease Control (*1*).

**Figure 1.**
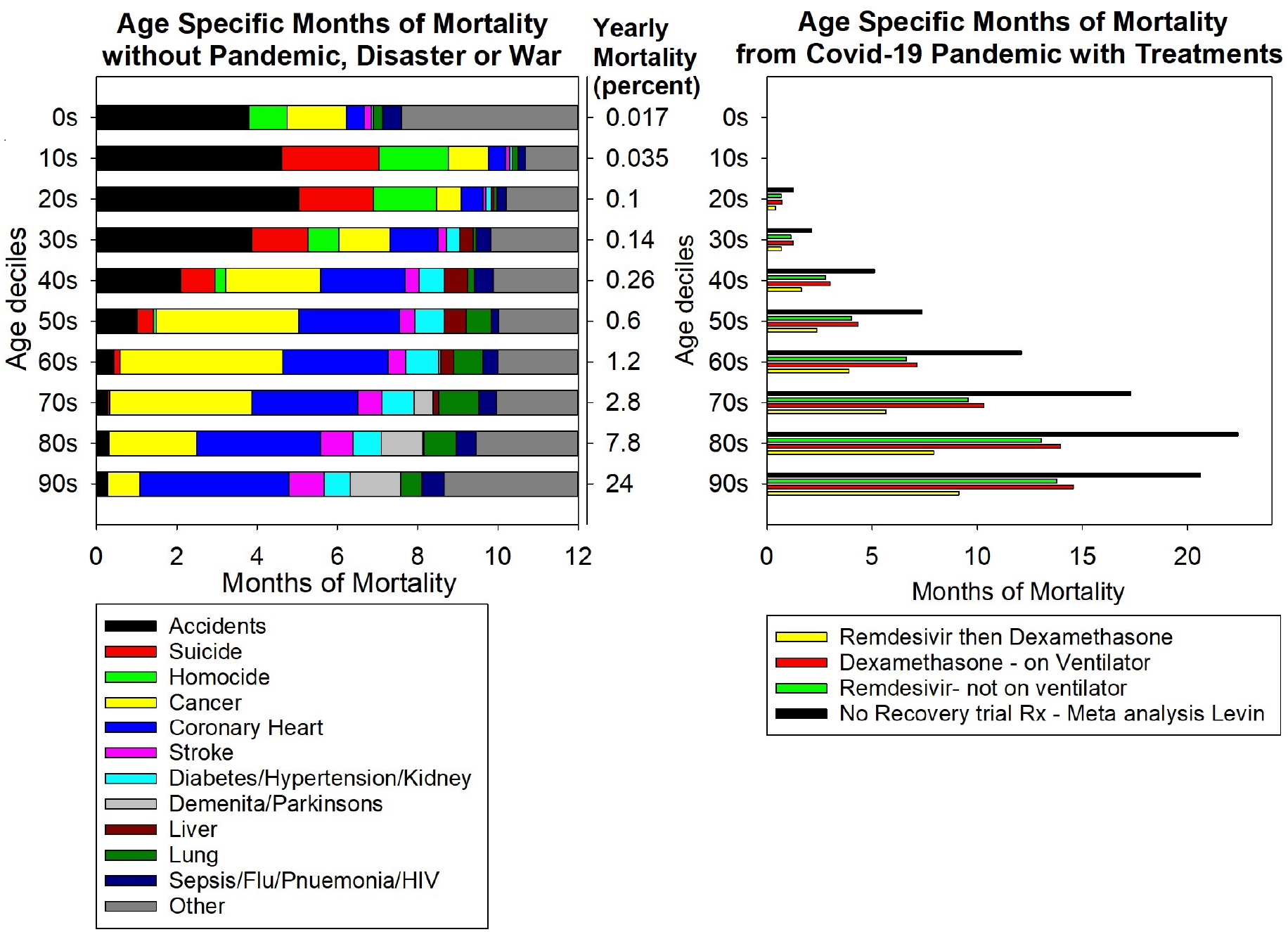
Age specific Months of Mortality (MOMa) from endemic (left) and one epidemic cause (Covid-19 in its first 6 months, right). The x axis scale is shrunk ∼twofold for covid-19.

Epidemic or pandemic diseases cause excess mortality that adds MOMa beyond 12 months. This is shown in the black bars on the right panel of Fig. 1 with data from a meta-analysis (*2*) of Covid-19 infection fatality rates in the winter and spring of 2020 (similar values were seen in three individual trials (*3-5*)). Also shown are the benefit from the Recovery trial of treating Covid-19 in non-ventilated people with remdesivir (*6*) (green bars), the benefit of treating Covid-19 in ventilated people with dexamethasone (*7*) (red bars), and the benefit of treating Covid-19 sequentially with remdesivir prior to ventilation and dexamethasone with ventilation (yellow bars, this assumes these treatments are additive).

For example, Fig. 1 shows that people in their 20s have a low risk of dying (0.1% yearly) and that the highest endemic risks are accidents (5 MOMa), followed by suicide (1.9 MOMa), homicide (1.6 MOMa) and cancer (1.0 MOMa). Covid-19 adds an excess 1.2 MOMa beyond endemic risk of dying. Treatment of Covid-19 with remdesivir (if not ventilated) or dexamethasone (if ventilated) based on the Recovery trial would reduce MOMa from 1.2 to 0.7.

People in their 40s also have a low risk of dying (0.26% yearly) and that the highest endemic risks are cancer (2.3 MOMa), coronary heart (2.1 MOMa), accidents (2.1 MOMa) and suicide (0.8 MOMa). Covid-19 adds an additional 5 MOMa beyond endemic risk of dying. Treatment of Covid-19 with remdesivir or dexamethasone would reduce MOMa from 5 to 3.

People in their 60s have an intermediate risk of dying (1.2% yearly) and that the highest endemic risks are cancer (4 MOMa), coronary heart (2.6 MOMa), diabetes/hypertension/kidney (0.8 MOMa) and lung (MOMa 0.7). Covid-19 adds an additional 12 MOMa beyond endemic risk of dying. Treatment of Covid-19 with remdesivir or dexamethasone would reduce MOMa from 12 to 7. Treatment of Covid-19 with remdesivir then dexamethasone sequentially would reduce MOMa from 12 to 4, a value similar to that year’s risk of dying from cancer.

People in their 80s have an high risk of dying (7.8% yearly) and that the highest endemic risks are coronary heart (3 MOMa), cancer (2.2 MOMa), dementia/parkinsons (1.0 MOMa), stroke (0.8 MOMa) and lung (0.8 MOMa). Covid-19 adds an additional 22 MOMa beyond endemic risk of dying. Treatment of Covid-19 with remdesivir or dexamethasone would reduce MOMa from 22 to ∼14. Treatment of Covid-19 with remdesivir then dexamethasone sequentially would reduce MOMa from 22 to 8, a value similar to that year’s risk of dying from coronary heart, cancer, dementia, parkinsons, stroke and lung combined.

The slope of the relation between MOMa and age decile shows how age affect mortality (Table 1). A negative slope suggests that cause of death predominates at younger ages and a positive slope suggests that cause of death predominates at older ages. A similar but likely less accurate measure is the decile associated with the highest MOMa.

**Table 1.** Two estimates of whether MOMa predominates at younger age (negative slope of MOMa vs age decile and lowest peak decile) or older age (positive slope of MOMa vs age decile and highest peak decile). MOMa values are those plotted in Fig. 1.

|  |  | Decile of age |  |  |  |  |  |  |  |  |  | Slope MOMa | Peak Decile |
| --- | --- | --- | --- | --- | --- | --- | --- | --- | --- | --- | --- | --- | --- |
|  |  | 90s | 80s | 70s | 60s | 50s | 40s | 30s | 20s | 10s | 0s |  |  |
|  | Yearly Percent Endemic Mortality | **24** | 7.8 | 2.8 | 1.2 | 0.6 | 0.26 | 0.14 | 0.1 | 0.035 | 0.017 | 0.47 | 90 |
| Age specific Months of Mortality (MOMa) |  |  |  |  |  |  |  |  |  |  |  |  |  |
| Endemic | Suicide |  |  | 0.04 | 0.16 | 0.41 | 0.86 | 1.39 | 1.86 | **2.43** |  | -0.40 | 10 |
|  | Homocide |  |  |  |  | 0.08 | 0.27 | 0.77 | 1.59 | **1.72** | 0.96 | -0.30 | 10 |
|  | Accidents | 0.29 | 0.31 | 0.28 | 0.43 | 1.01 | 2.09 | 3.87 | **5.03** | 4.61 | 3.79 | -0.27 | 20 |
|  | Liver |  | 0.03 | 0.13 | 0.32 | 0.55 | **0.58** | 0.33 | 0.06 |  |  | -0.09 | 40 |
|  | Other | 3.35 | 2.55 | 2.05 | 1.99 | 1.98 | 2.12 | 2.17 | 1.80 | 1.31 | **4.40** | 0.00 | 0 |
|  | Sepsis/Flu/Pnuemonia/HIV | **0.55** | 0.49 | 0.44 | 0.38 | 0.18 | 0.47 | 0.36 | 0.23 | 0.18 | 0.48 | 0.06 | 90 |
|  | Cancer | 0.80 | 2.18 | 3.55 | **4.06** | 3.53 | 2.35 | 1.27 | 0.60 | 1.00 | 1.49 | 0.08 | 60 |
|  | Diabetes/Hypertension/Kidney | 0.65 | 0.71 | 0.80 | **0.83** | 0.72 | 0.63 | 0.34 | 0.14 | 0.07 | 0.06 | 0.18 | 60 |
|  | Coronary Heart | **3.71** | 3.09 | 2.64 | 2.61 | 2.52 | 2.12 | 1.20 | 0.54 | 0.42 | 0.44 | 0.20 | 90 |
|  | Lung | 0.52 | 0.81 | **1.00** | 0.74 | 0.64 | 0.18 | 0.08 | 0.07 | 0.15 | 0.23 | 0.20 | 70 |
|  | Stroke | **0.87** | 0.80 | 0.60 | 0.44 | 0.38 | 0.35 | 0.21 | 0.08 | 0.10 | 0.16 | 0.22 | 90 |
|  | Demenita/Parkinsons | **1.26** | 1.02 | 0.47 | 0.05 |  |  |  |  |  |  | 0.60 | 90 |
| Pandemic | Covid 19 Meta analysis (Levin) | 20.61 | **22.38** | 17.28 | 12.09 | 7.35 | 5.09 | 2.09 | 1.23 |  |  | 0.30 | 80 |

|  |  | Decile of age |  |  |  |  |  |  |  |  |  | Slope | Peak |
| --- | --- | --- | --- | --- | --- | --- | --- | --- | --- | --- | --- | --- | --- |
|  |  | 90s | 80s | 70s | 60s | 50s | 40s | 30s | 20s | 10s | 0s | MOMa | Decile |
|  | Yearly Percent Endemic Mortality | **24** | 7.8 | 2.8 | 1.2 | 0.6 | 0.26 | 0.14 | 0.1 | 0.035 | 0.017 | 0.47 | 90 |
| Age specific Months of Mortality (MOMa) |  |  |  |  |  |  |  |  |  |  |  |  |  |
| Endemic | Suicide |  |  | 0.04 | 0.16 | 0.41 | 0.86 | 1.39 | 1.86 | **2.43** |  | -0.40 | 10 |
|  | Homocide |  |  |  |  | 0.08 | 0.27 | 0.77 | 1.59 | **1.72** | 0.96 | -0.30 | 10 |
|  | Accidents | 0.29 | 0.31 | 0.28 | 0.43 | 1.01 | 2.09 | 3.87 | **5.03** | 4.61 | 3.79 | -0.27 | 20 |
|  | Liver |  | 0.03 | 0.13 | 0.32 | 0.55 | **0.58** | 0.33 | 0.06 |  |  | -0.09 | 40 |
|  | Other | 3.35 | 2.55 | 2.05 | 1.99 | 1.98 | 2.12 | 2.17 | 1.80 | 1.31 | **4.40** | 0.00 | 0 |
|  | Sepsis/Flu/Pnuemonia/HIV | **0.55** | 0.49 | 0.44 | 0.38 | 0.18 | 0.47 | 0.36 | 0.23 | 0.18 | 0.48 | 0.06 | 90 |
|  | Cancer | 0.80 | 2.18 | 3.55 | **4.06** | 3.53 | 2.35 | 1.27 | 0.60 | 1.00 | 1.49 | 0.08 | 60 |
|  | Diabetes/Hypertension/Kidney | 0.65 | 0.71 | 0.80 | **0.83** | 0.72 | 0.63 | 0.34 | 0.14 | 0.07 | 0.06 | 0.18 | 60 |
|  | Coronary Heart | **3.71** | 3.09 | 2.64 | 2.61 | 2.52 | 2.12 | 1.20 | 0.54 | 0.42 | 0.44 | 0.20 | 90 |
|  | Lung | 0.52 | 0.81 | **1.00** | 0.74 | 0.64 | 0.18 | 0.08 | 0.07 | 0.15 | 0.23 | 0.20 | 70 |
|  | Stroke | **0.87** | 0.80 | 0.60 | 0.44 | 0.38 | 0.35 | 0.21 | 0.08 | 0.10 | 0.16 | 0.22 | 90 |
|  | Demenita/Parkinsons | **1.26** | 1.02 | 0.47 | 0.05 |  |  |  |  |  |  | 0.60 | 90 |
| Pandemic | Covid 19 Meta analysis (Levin) | 20.61 | **22.38** | 17.28 | 12.09 | 7.35 | 5.09 | 2.09 | 1.23 |  |  | 0.30 | 80 |

For example, some causes of death predominate in younger people: accidents’ slope is −0.27 and a peak decile in the 20s, suicide’s slope is −0.40 and a peak decile in the 10s, and homicide’s slope is −0.30 and a peak decile in the 10s. Other causes of death predominate in middle age: liver’s slope is −0.09 and a peak decile in the 40s, and cancer’s slope is +0.08 and a peak decile in the 60s. Other causes of death predominate at older ages: coronary heart’s slope is +0.20 and a peak decile in the 90s and Covid-19’s slope is −0.30 and a peak decile in the 80s.

## DISCUSSION

This method allows people in various deciles of life to ascertain which causes of death are more and less likely for their age. It also makes these risks apparent to policy makers and physicians. For example, even during the covid-19 pandemic, people from age 10-39 are at most risk from accidents, suicide and homicide, so public health measures should concentrate on these risks in addition to pandemic issues such as younger people transmitting covid-19 to others. During the pandemic, people in their 60s are at most risk from covid-19 and cancer.

The endemic MOMa are based only on the year 2015 in the US, but are based on large numbers and so are likely not to change much over a decade. However, they will likely change over longer periods. These MOMa are also likely representative of other higher income countries with the exception of homicide which varies by country. They should be recalculated for medium and lower income countries and when new therapies change causes of death. For example, the fall in blood pressure with treatment over the last 40 years (*8*) is likely responsible for a significant part of the decline in coronary heart, stroke and renal disease deaths over this time. Even more benefit over the last two centuries has occurred with clean water reducing diarrheal deaths from diseases such as cholera.

The MOMa for covid-19 are based on data in the first 6 months of the pandemic. Some reports suggest than the covid-19 mortality has fallen dramatically (*9*), possibly from improvement in treatment and/or infection of different populations. Lower covid-19 mortality would reduce the MOMa values.

This analysis includes only mortality and not morbidity which can be substantial from any of these diseases. A similar statistic could also be calculated for any specific morbidities.

Months of mortality can also be stratified by Sex (MOMs), race (MOMr) or any other subgroup.

## CALCULATION METHODS

See attached excel file for calculations: all calculations are included.

Probabilities of different outcomes are additive. So the percent of deaths (total or specific cause) in each age group is calculated directly:

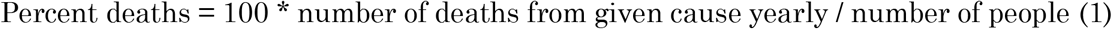

Months of mortality for a specific cause and age decile (MOMa) is calculated as:

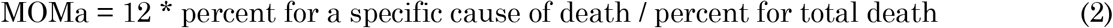

When data are presented in different age ranges, then probabilities needed to be interpolated. Unfortunately, changes in probability are not additive. Changes in probability are additive once data are transformed into a linear probability space (*10, 11*). For example, if there are percentage data for 35-44 and 45-54 year olds, then to average to get 40-49 year olds, calculation is done as follows:

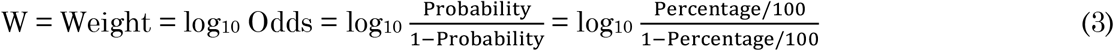

W35-44 year old and W45-54 year old are calculated and then added and transformed back (multiply probability by 100 to get percentage:

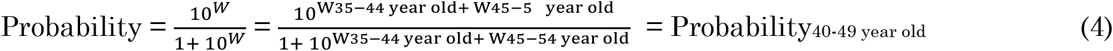

For treatments Bayes theorem in a linear probability space (*10, 11*) is:

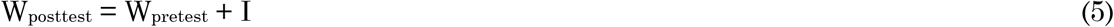

I stands for impact which is an estimate of benefit or harm of a test or treatment. I is similar to effect size and is calculated as the difference of W values with and without treatment in a clinical trial.

See publications (*10, 11*) for details on how this system of W (weights, an estimate of probability) and I values (impact, an estimate of effect size) can allow easy calculation of Bayes theorem.

## Data Availability

data (one xls) will be presented when published

